# Optimizing Dose-Response Decisions in Psoriatic Arthritis via Causal Machine Learning: A Real-World Evaluation of Secukinumab Treatment

**DOI:** 10.1101/2025.10.11.25335913

**Authors:** Uta Kiltz, Thomas Glassen, Jan Brandt-Juergens, Ferenc Kiss, Daniel Peterlik, Benjamin Gmeiner

**Author notes:** These authors contributed equally (Uta Kiltz, Thomas Glassen).

## Abstract

Personalized treatment in psoriatic arthritis (PsA) remains challenging, particularly in guiding dose escalation decisions. We applied a causal machine learning framework to real-world data from the AQUILA study to evaluate the impact of secukinumab dose escalation (150 mg to 300 mg) on disease activity and health-related quality of life (HR-QoL). Using double machine learning, we estimated both average and patient-level conditional treatment effects (CATEs) based on 42 baseline variables, including demographics, laboratory values, disease activity, HR-QoL, comorbidities, and prior treatments.

Patients receiving the higher dose showed greater improvement in Psoriatic Arthritis Impact of Disease (PsAID) scores compared to the lower dose (mean reduction: 1.81 vs. 1.44). Subgroup analysis revealed a 28% HR-QoL gain in patients with elevated body mass index (BMI) or C-reactive protein (CRP). The model predicted that approximately 75% of patients would benefit from dose escalation.

These findings demonstrate the utility and scalability of causal machine learning for quantifying individualized treatment effects and guiding personalized treatment decisions in PsA. The approach is transferable to other chronic conditions, supporting more precise, data-driven care in real-world clinical settings.

## INTRODUCTION

The promise of personalized medicine lies in matching the right treatment - and dose - to the right patient. In patients with psoriatic arthritis (PsA), however, clinicians often rely on judgement rather than data when deciding on dose escalation. This gap reflects a broader challenge: how to identify treatment effect heterogeneity using real-world data.

PsA is a chronic heterogeneous inflammatory immune-mediated disease characterized by inflammation of the musculoskeletal system, including arthritis, enthesitis, dactylitis, and spondylitis^1,2^. Approximately 30% of patients with psoriasis may develop PsA, especially those with severe psoriasis or involvement of the nails or scalp^2-4^. The prevalence of PsA in the general population ranges from 0.1% to 1%^2,5,6^, and the incidence is estimated to be 83 per 100,000 person-years (PY) [95% confidence interval (CI), 41–167 per 100,000 PY]^2,7^. The use of conventional synthetic disease-modifying antirheumatic drugs (csDMARDs) is recommended as the first-line treatment for patients with PsA in treatment guidelines, followed by the use of biological disease-modifying antirheumatic drugs (bDMARDs), apremilast, or targeted synthetic DMARDs (tsDMARDs)^8^. Recent advancements have led to the development of targeted therapies for psoriatic disease on the basis of the role of IL-23/IL-17^2,8-10^.

Secukinumab, a fully human monoclonal antibody that selectively targets IL-17A, is approved for the treatment of patients with PsA. Secukinumab has shown superior efficacy to placebo in PsA patients with arthritis, spondylitis, dactylitis, enthesitis, and skin and nail disease^11-15^. The European Medicines Agency (EMA) has approved an updated label for secukinumab, allowing an increase in dose to 300 mg from 150 mg in adults with active radiographic axial spondyloarthritis (r-axSpA) or active PsA when a treatment response is not achieved^16^. This update provides clinicians with more treatment options to achieve better patient responses and can lead to better treatment outcomes and potentially reduce disease progression. In clinical practice, physicians typically follow the approval status and recommend a higher dose for patients with prior biologic experience, moderate to severe PsA symptoms, or body weight exceeding 90 kilograms^16^. However, adherence to these guidelines is inconsistent in clinical practice, and dose decisions are often based on the clinical judgement of the individual physician.

Evidence suggests that increasing the dose of secukinumab from 150 mg to 300 mg can improve efficacy outcomes in PsA patients^14,17^. A phase III study also demonstrated greater improvements in psoriasis symptoms with a higher dose of secukinumab^12^. Although there is evidence of improved efficacy with secukinumab dose escalation in patients with PsA, details on how much treatment escalation can improve outcomes within subgroups of patients or even at an individual level remain unclear.

To overcome the shortcomings of guiding evidence, the use of causal machine learning (ML)-based models can be explored.

While supervised ML methods have been applied to predict treatment outcomes in patients with PsA/ankylosing spondylitis (AS)^18^, to our knowledge, causal ML techniques have not yet been applied to evaluate the effects of updosing in PsA.

In this context, causal inference plays a vital role in bridging the gap between prediction and decision-making. Causal inference is a systematic approach for comprehending the cause-and-effect relationships between variables or events. By identifying and understanding the factors that contribute to a specific outcome, causal inference provides valuable insights. This method is widely employed in fields such as social sciences, economics, and clinical research. In clinical research, causal analysis has proven valuable in analysing data from clinical trials^19^, especially when the overall population has a poor response or a weak or no response to a particular treatment. Through this method, researchers can assess treatment effect heterogeneity, providing a deeper understanding of the factors that influence treatment outcomes and identifying subgroups that respond more favourably to treatment^20^.

## OBJECTIVE

The primary objective was to develop a causal ML model that is based on data from the AQUILA non-interventional study. This model aims to predict the impact of a hypothetical increase in secukinumab dose from 150 mg (low) to 300 mg (high) on (i) disease activity and (ii) health-related quality of life (HR-QoL) in patients with PsA. The model offers insights into which patient subgroups are most likely to benefit from dose escalation.

## RESULTS

Data from 1994 PsA patients were used in this ML study, and 1235 fulfilled the inclusion criteria for the analysis (Figure 1). The baseline demographics and characteristics of the patients are presented in Table 1.

**Table 1.** Baseline demographics and characteristics of patients.

| Characteristics | Low dose (150 mg, n = 616) |  | High dose (300 mg, n = 619) |  |
| --- | --- | --- | --- | --- |
| | Count | Mean ± SD | Count | Mean ± SD |
| Age | 616 | 53.3 ± 11.3 | 619 | 52.3 ± 11.7 |
| Female, n (%) | 616 | 377 (61.2) | 619 | 335 (54.1) |
| BMI | 591 | 28.8 ± 5.7 | 608 | 30.0 ± 6.5 |
| CRP level (mg/L) | 476 | 8.4 ± 14.2 | 497 | 8.3 ± 13.2 |
| PASI, 0–72 | 216 | 4.7 ± 8.6 | 279 | 9.4 ± 12.3 |
| WJC, 0–67.3 | 392 | 5.7 ± 6.3 | 406 | 7.1 ± 7.2 |
| SJC, 0–66 | 393 | 3.6 ± 4.5 | 406 | 4.4 ± 6.2 |
| TJC, 0–68 | 393 | 6.8 ± 8.2 | 406 | 8.4 ± 8.5 |
| PsAID score, 0–10 | 616 | 5.1 ± 2.1 | 619 | 5.4 ± 2.1 |
| PGA score, 0–10 | 458 | 5.3 ± 2.5 | 432 | 5.3 ± 2.5 |
| BDI score, 0–63 | 545 | 12.8 ± 9.4 | 544 | 12.6 ± 9.6 |
| MOS Sleep score, 0.9–5.9 | 378 | 3.4 ± 0.4 | 383 | 3.4 ± 0.4 |
| PhGA score, 0–10 | 560 | 5.5 ± 2.0 | 574 | 5.6 ± 2.1 |
| SPI-II score, 16.8–65.9 | 356 | 42.6 ± 9.3 | 367 | 42.4 ± 10.1 |

Abbreviations: BDI: beck depression inventory; BMI: body mass index; CRP: C-reactive protein; MOS: medical outcomes study sleep scale; PASI: psoriasis area and severity index; PGA: patient global assessment; PhGA: physician's global assessment; PsAID: psoriatic arthritis impact of disease; SJC: swollen joint count; SPI-II: sleep problems index-II; TJC: tender joint count; WJC: weighted joint count.
Notes: Only data from patients with PsAID scores at baseline and visit 3 or visit 4 are presented.

**Figure 1:**
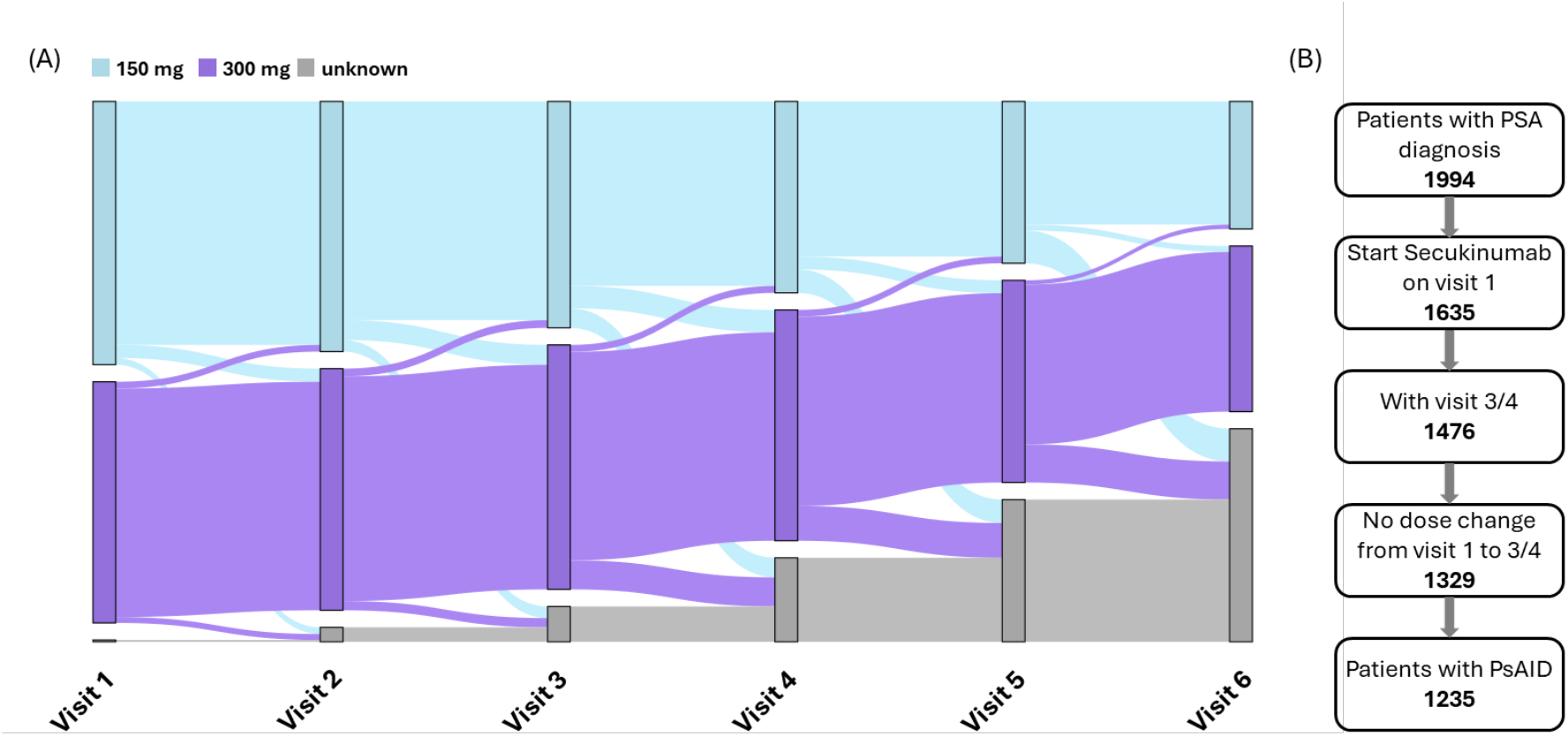
(**a**) Sankey plot showing patient adherence in the 150 mg and 300 mg dose groups and (**b**) flow chart of the attrition of patients from PsAID assessment in the AQUILA study. The right panel displays patient dropout from PsAID assessments. The left panel depicts changes in treatment status throughout the six visits. The length of each bar and the link between bars are proportional to the number of patients. While slightly more patients began at a low dose, over the course of the visits, more patients transitioned to a high dose than did those who switched to a low dose. Ultimately, more patients were on the high dose. PsAID, Psoriatic Arthritis Impact of Disease. Visits were scheduled at baseline (Visit 1 at week 0) and at weeks 4, 16, 28, 40, and 52.

Among the 1235 patients, 616 received 150 mg secuki-numab (low dose), and 619 received 300 mg secukinumab (high dose) at the baseline visit. In general, the high-dose group presented numerically higher disease activity scores and HR-QoL scores at baseline, indicating more severe disease symptoms (see Table 1). The relevant demographics were similar between the two groups.

### Disease activity and HR-QoL outcomes

Changes in disease activity scores and HR-QoL scores are illustrated in Figure 2. The number of missing values varied between 76% for the PASI and 38% for the PsAID.

**Figure 2:**
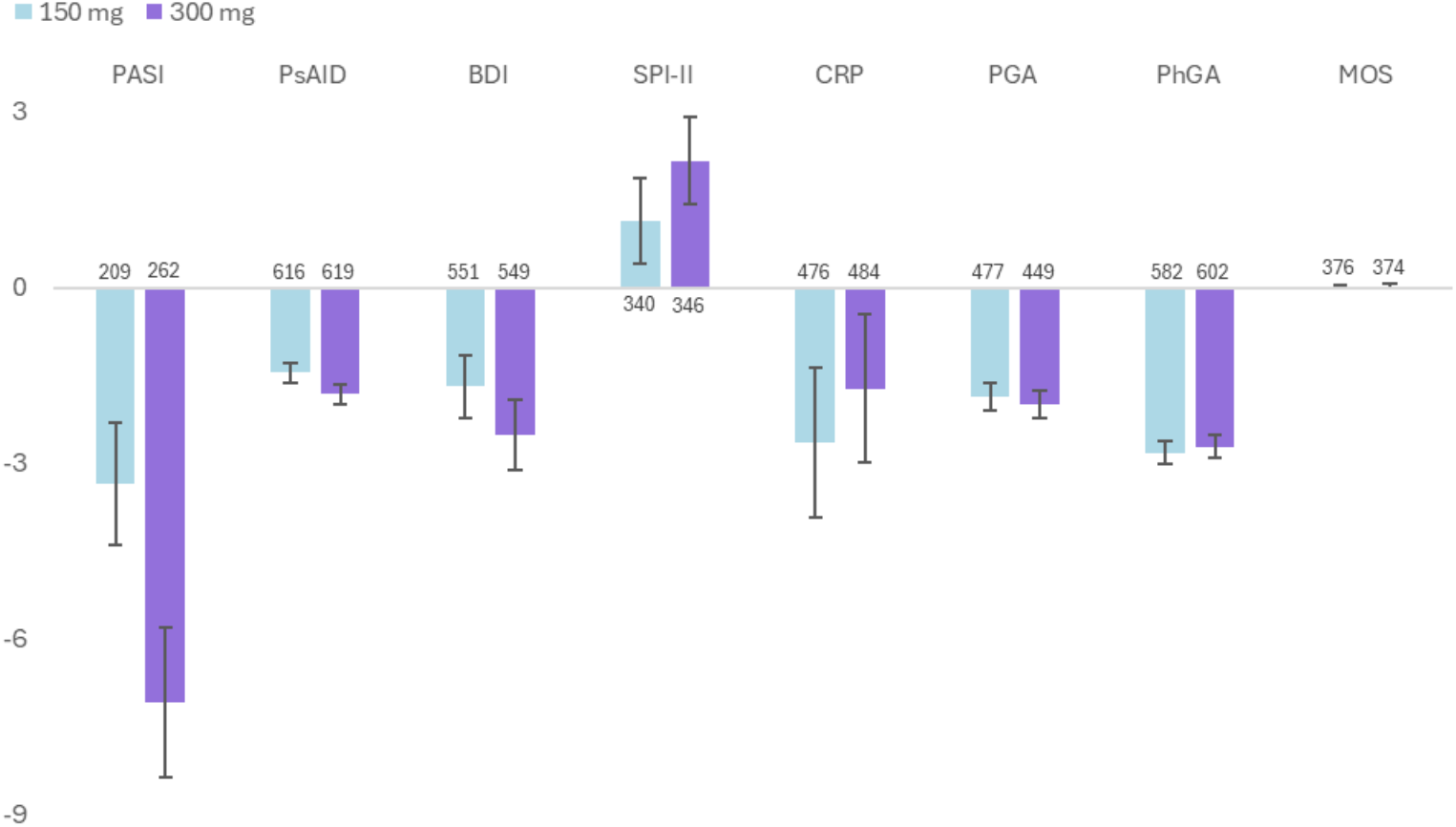
Observed disease activity and HR-QoL outcome changes for 150 and 300 mg doses at weeks 28 or 40 in the AQUILA study. The case numbers for each dose and dimension are shown near the axis. To show the most important differences first, dimensions are ordered by the relative overlap of their dose-specific 95% confidence intervals, from least to most. BDI-II, Beck Depression Inventory-II; PASI, Psoriasis Area and Severity Index; MOS, Medical Outcomes Sleep Scale; PhGA, Physician’s Global Assessment; PGA, Patient Global Assessment; PsAID, Psoriasis Arthritis Impact of Disease; SPI-II, Sleep Problems Index-II; CRP, C-reactive protein.

Significant changes in scores between baseline and Visit 3/4 (at week 16-28) were observed for the PASI (*p* < 0.001), PsAID (*p* < 0.01), and BDI (*p* < 0.05). The PsAID score was selected as the primary outcome measure because of the high completeness of the data.

The high-dose cohort had a numerically greater (*p* > 0.05) baseline PsAID score of 5.35 than did the low-dose cohort (5.12) (Table 1 and Figure 3(c)).

**Figure 3:**
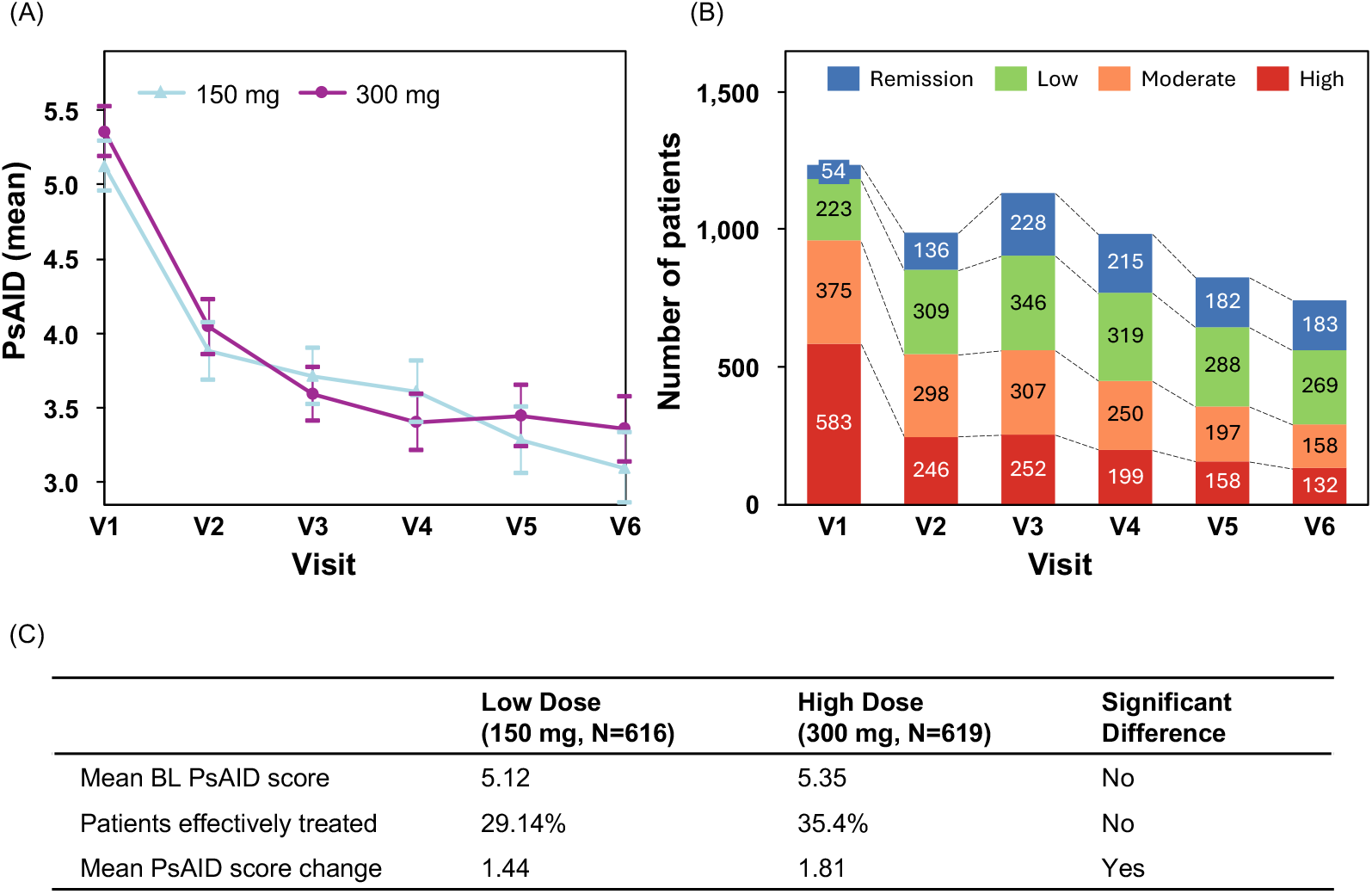
Observed HR-QoL (PsAID) across visits. (**a**) Line plot showing the PsAID scores of patients receiving low (150 mg) and high (300 mg) secukinumab doses across six visits. (**b**) Patient distribution into high, moderate, and low HR-QoL and remission categories over six visits. (**c**) Table comparing low- and high-dose groups: mean baseline PsAID score, percentage of effectively treated patients, and mean PsAID score change. HR-QoL, health-related quality of life; PsAID, Psoriatic Arthritis Impact of Disease.

The average PsAID score reduction after treatment was -1.63 points. While the overall population achieved a PsAID score reduction, the high-dose cohort had a greater reduction (*p* < 0.001) of 1.81 PsAID points compared with 1.44 PSAID points for the low-dose cohort at Visit 3.

When effective treatment was defined as a PsAID score change of ≥3 points, 29.1% and 35.4% of patients achieved this under low and high doses, respectively (Figure 3(c)). From the curvature of the line plot for both doses (Figure 3(a)), most changes in value occurred between Visit 1 and Visit 3, and after Visit 3, the line was much flatter; therefore, the change between Visits 1 and 3 was used as the target value for causal analysis.

The pooled distribution of patients across HR-QoL categories (remission, low, moderate, high impact) over time is shown in Figure 3b, indicating a general shift toward better health-related quality of life between Visit 1 and Visit 3. The PsAID score distribution at baseline and PsAID score changes after 16-28 weeks are presented in Supplementary Figure 1.

### Model features

Fitting the causal model to the full cohort of 1235 patients revealed a global causal effect for each feature. The global causal effect represents the overall effect considering all patients, as opposed to local effects that focus on specific individual or subgroup responses. The associated coefficients with the respective *p* values are depicted in the forest plot in Figure 4.

**Figure 4:**
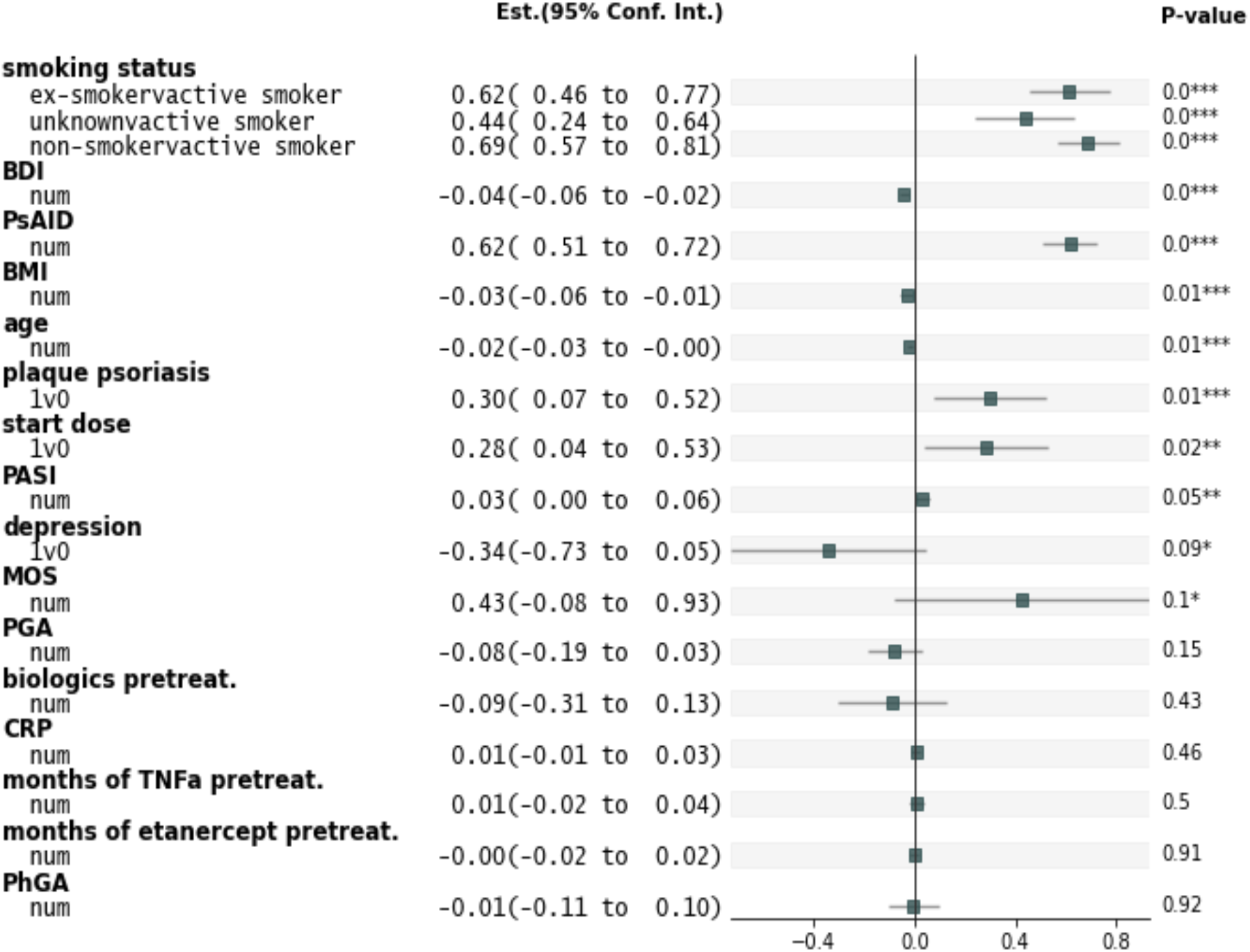
Forest plot of model coefficients and *p* values. Coefficients are sorted by *p* value in ascending order. Each coefficient represents the PsAID score change per unit feature change. For example, a start dose change from 0 (150 mg) to 1 (300 mg) increased the PsAID score by 0.28 points. BDI, Beck Depression Inventory; BMI, body mass index; CRP, C-reactive protein; PASI, Psoriasis Area and Severity Index; PhGA, Physician’s Global Assessment; MOS, Medical Outcomes Study Sleep Scale; PGA, Patient Global Assessment; PsAID, Psoriatic Arthritis Impact of Disease; biologics pretreat., number of pretreatments with biologics; months of etanercept pretreat., months pretreated with etanercept; smoking status, smoking status with active smoker as baseline, e.g., ex-smokervactive smoker depicts the relative effect of ex-smoker status against active smoker status on the PsAID score; 1v0, binary (yes/no); num, numerical.

### Response to treatment in subgroups

The causal model yielded a significant *p* value for “Start dose,” indicating a significant effect of updosing on HR-QoL, with the true average treatment effect (ATE) estimated at 0.236 units (compared with the biased 0.37 units).

The heterogeneity tree in Figure 5 shows modelled sub-group changes in response differences in the PsAID between the 150 mg and 300 mg doses. The root node CATE of 0.24 corresponds to the ATE, as there are not yet any conditions applied. A total of 918 patients with a BMI ≤ 32.5 exhibited a poor response difference between treatments, with a CATE of 0.14, whereas 317 patients with a higher BMI had a more favourable CATE of 0.52. Further divisions can be made for more precise treatment response differences, as shown in Figure 5. Notably, 613 patients within the leaves (lowest level nodes) with green boundaries reduced their PsAID score on average by 0.45 points with updosing from 150 to 300 mg, a 28% improvement over the status quo

**Figure 5:**
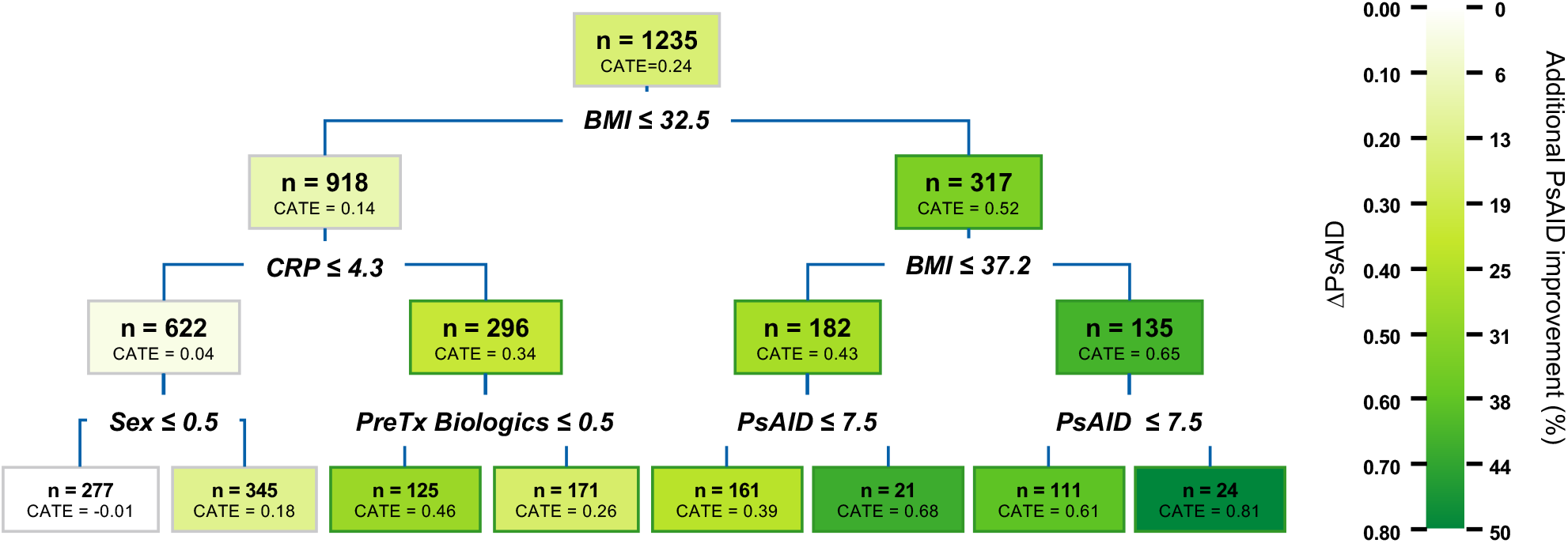
Predicted heterogeneity tree illustrating subgroup treatment responses. The heterogeneity tree resembles a decision tree, but it splits at points that maximize heterogeneity in the resulting “child” nodes. Each leaf represents a subgroup of samples whose response to treatment is distinct from that of the other samples. The root node indicates the entire population with an ATE difference of 0.236 units, whereas nodes with green boundaries represent subgroups that respond more favourably to the treatment, showing a 28% improvement versus 15% for the overall population. ATE, average treatment effect; CATE, conditional average treatment effect difference; PsAID, Psoriatic Arthritis Impact of Disease; preTx Biologics, number of pretreatments with biologics; CRP, C-reactive protein; BMI, body mass index.

### Response to treatment at the individual level

The distribution of predicted treatment effects on PsAID scores among patients receiving low and high doses if they hypothetically switched to the other dose group is shown in Figure 6.

**Figure 6:**
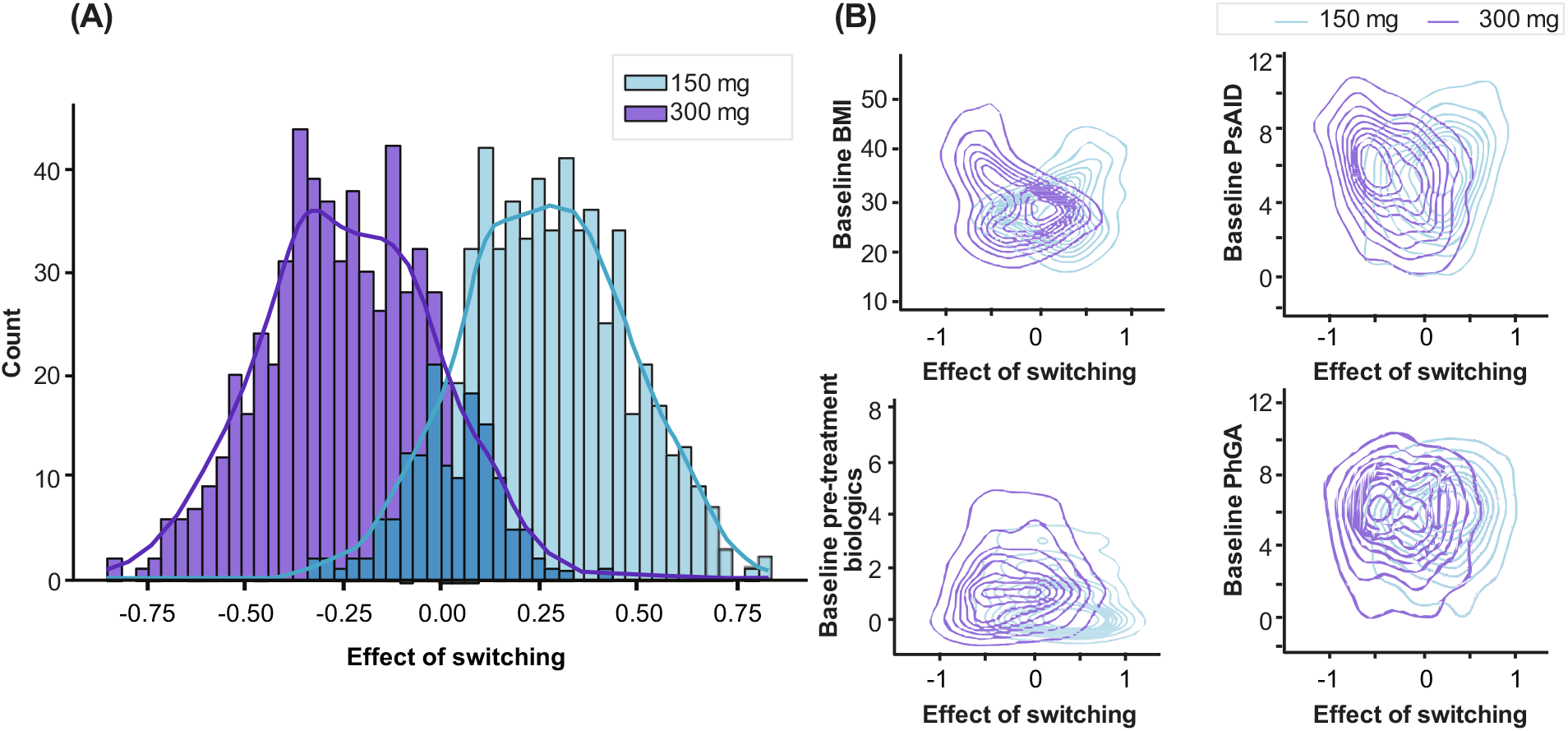
Predicted overall impact of a hypothetical initial treatment switch on PsAID scores. (a) Histogram: Approximately 75% of patients who switched from the low to the high dose could achieve up to a 0.8-point gain in benefit, whereas 75% of patients who switched from the high to the low dose might lose up to 0.8 points of benefit. (**b**) Covariate impact 2D density plots: High effectiveness for patients with “BMI” > 38 when switching doses is shown at the top left, whereas for “pretreatment biologics count”, no distinct dose−response difference is visible in the lower left plot. Low dose: 150 mg; high dose: 300 mg. PsAID, Psoriatic Arthritis Impact of Disease; BMI, body mass index.

To provide another perspective on treatment effects, we assessed patient shifts between predefined burden categories following a simulated treatment switch.

The outcomes for the low-dose cohort (top row) and the corresponding distribution of potential outcomes after dose escalation (bottom row) are shown in Figure 7.

**Figure 7:**
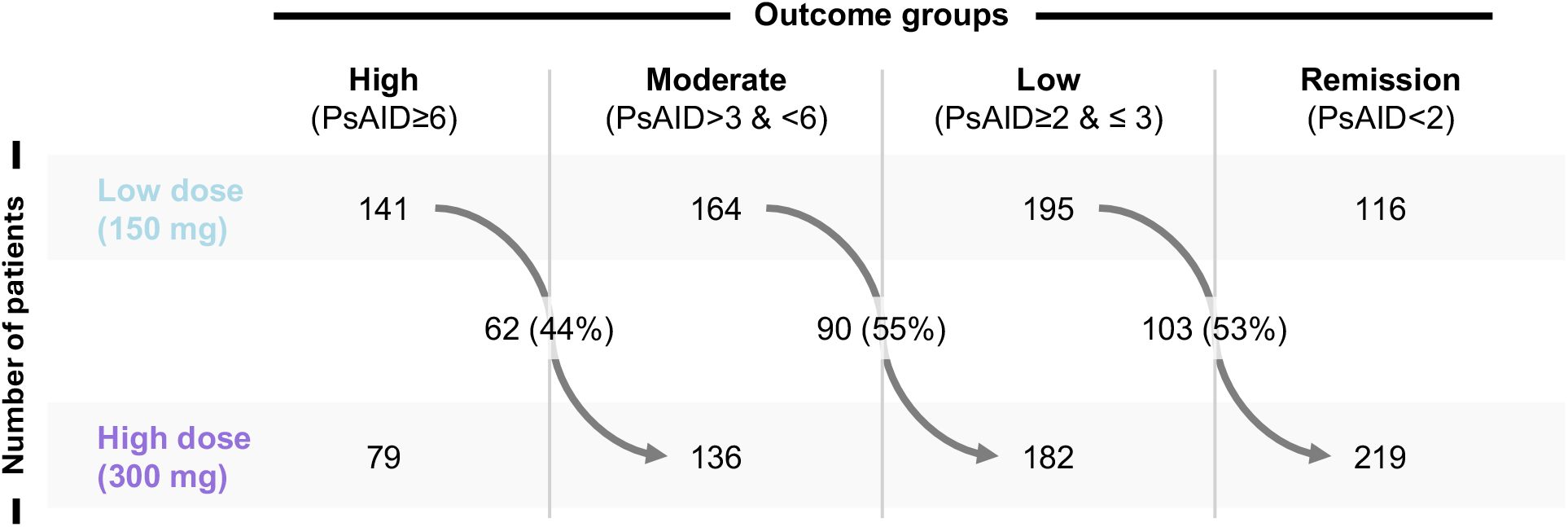
Predicted patient burden category shifts after dose increases considering the counterfactual outcomes. The top row shows the distribution of patient outcomes under low-dose treatment, whereas the bottom row represents potential outcome distributions after dose escalation. The arrows indicate the absolute number of patients shifting between categories, with percentages shown in parentheses.

Sixty-two out of 141 high-burden patients (44%) transitioned to the moderate burden category. Among the 164 patients with a moderate burden at the low dose, 90 (55%) shifted to the low burden category. Similarly, of the 195 patients classified as having a low burden under the low dose, 103 (53%) moved into remission after the dose increase.

## DISCUSSION AND OUTLOOK

In clinical studies, most PsA patients experience a substantial decrease in disease activity during the initial three months of biologic treatment. This observation guided us to focus our analysis on the period from Visit 1 to Visit 3/4. During this timeframe, we evaluated changes in 6 HR-QoL and 3 disease activity measures in PsA patients. Among these, only changes in PsAID scores at baseline were significantly different between the two cohorts.

Notably, the high-dose cohort had a higher baseline PsAID score (5.35) than did the low-dose cohort (5.12). Both groups demonstrated improvements in PsAID scores, with the high-dose group showing a more pronounced reduction (1.81 vs. 1.44) over approximately 16 weeks. While the low-dose cohort appeared to be more effectively treated in terms of PhGA scores, this difference was numerical rather than statistically significant.

Our causal ML model suggests that increasing the dose of secukinumab could significantly improve treatment outcomes for PsA patients. The model predicts that approximately 75% of all patients should receive a higher dose, and 75% of those currently on one dose should switch. However, these findings are based solely on model predictions and do not account for the current approval status of secukinumab, emphasizing the need for regulatory considerations before implementing dose changes in clinical practice.

Given real-world treatment, assignment is often subject to selection bias. Patients were not randomly allocated to the groups, and those in the 300 mg dose cohort typically presented with more severe symptoms. Hence, the observed PsAID score difference between the low- and high-dose cohorts is likely influenced by selection bias. To address this, we developed a causal model employing methods such as inverse probability of treatment weighting (IPTW) to mitigate bias.

Although the significant effect of updosing on HR-QoL, with the true ATE estimated at 0.236 units (compared with the biased 0.37 units), may appear modest, it represents a notable improvement compared with the average reduction of approximately -1.63 units in the low-dose cohort, signifying a 15% improvement.

The causal model identified several significant features, including “smoking status”, “PsAID”, “BMI”, “BDI”, and “plaque psoriasis”. Patients with high baseline PsAID scores usually start with higher doses, which significantly affects PsAID score changes. Baseline PsAID scores also moderately correlate with BDI scores, which might explain why the change in BDI score was identified as significant. Additionally, smokers responded less favourably to low doses than to high doses, underscoring the need for higher doses because of the detrimental impact of smoking on the treatment response (see Supplementary Figure 2)^21^. Moreover, BMI aligns with guidelines suggesting high doses for patients exceeding 90 kg, whereas plaque psoriasis often indicates more severe PsA and thus a higher warranted dose. Additionally, EMA guidelines recommend a 300 mg dose for TNF inhibitor (TNFi)-inadequate responders.

The causal model can generate a straightforward decision tree to determine optimal doses for patients. For example, patients with an elevated BMI or high CRP level should receive high doses, leading to outcome improvements ranging from 15% to 28% among these groups.

The purpose of using supervised ML algorithms is to minimize discrepancies between actual and predicted values. While these models can be highly accurate, they may not always yield the desired results when applied in practice. Accurate models may also lack sufficient guidance for system modifications or decision-making. A notable limitation of the model is its failure to account for drug costs and potential side effects. Higher doses tend to be recommended on the basis of model predictions, given the superior treatment effects of these doses. However, lower doses are more cost-effective, making them preferable unless the higher dose substantially improves treatment efficacy (e.g., a PsAID score reduction of more than 0.2 points). Without a defined threshold, 98% of patients would commence treatment on a higher dose; with a 0.2 point threshold, 66% should start treatment on a higher dose; and with a 0.5 point threshold, only 9% of patients should begin treatment on a higher dose. Despite these limitations, this study demonstrates the feasibility and value of applying causal ML to real-world data to inform personalized dosing strategies in PsA patients. By identifying subgroups most likely to benefit from increased doses, our model can serve as a robust tool for personalized treatment strategies, optimizing clinical outcomes in a cost-effective manner. This approach highlights the need for a nuanced understanding of dose−response relationships and the potential for personalized medicine to improve patient care. The aim of future studies should be to refine these models further, incorporating additional variables such as drug costs and side effect profiles, to develop even more comprehensive treatment guidelines. Ultimately, integrating causal ML into clinical practice holds promise for transforming the management of PsA and other complex diseases, leading to improved patient outcomes and more efficient health care delivery.

Moreover, it is worth noting the potential for such models to be extended beyond PsA to other diseases in which tailored treatment plans are required. The ability of these models to account for individual patient characteristics and predict optimal interventions underscores their significance. The continuous refinement of these models, alongside advancements in data collection and analysis techniques, will be pivotal in their successful implementation. It is essential to engage healthcare professionals, researchers, and stakeholders in a collaborative effort to harness the full potential of causal ML, ensuring that the benefits are maximized for patients and healthcare systems alike.

## METHODS

### AQUILA study

AQUILA is an ongoing multicentre, prospective, non-interventional study designed to assess the effectiveness, safety, and adherence rates of secukinumab in patients with active PsA or r-axSpA in routine clinical practice in Germany^22^. The study includes patients aged ≥18 years with high disease activity who are either receiving secukinumab or scheduled to initiate treatment based on therapeutic need. Patients were treated following the approved guidelines, initially receiving either 150 mg or an increased 300 mg dose of secukinumab, with the dose being adjusted as needed throughout the study. Exclusion criteria were data unavailability for the 52-week observation period, concurrent participation in other clinical or non-interventional studies involving secukinumab, or existing contraindications to secukinumab. Each patient was observed for a planned period of 52 weeks^22^. Data was collected over the observational period, with visits scheduled at baseline (week 0) and at weeks 4, 16, 28, 40, and 52. The data collected for assessing disease activity and HR-QoL in patients with PsA included scores for the Psoriatic Arthritis Impact of Disease (PsAID), Beck Depression Inventory (BDI), Medical Outcomes Study Sleep Scale (MOS Sleep), Physician’s Global Assessment (PhGA), Sleep Problems Index-II (SPI-II), Psoriasis Area and Severity Index (PASI) response, and visual analogue scale (VAS) for global disease activity, global pain, and nighttime pain.

### Patient selection

Patients were included if they were adults (aged ≥18 years) with a physician-confirmed diagnosis of active PsA who started secukinumab treatment not earlier than 4 weeks before the baseline. Additionally, patients needed to have records available at both baseline and Visit 3 or, if Visit 3 data were missing, Visit 4. During this period, patient secukinumab dose had to remain unchanged. To ensure consistency in the analysis, only patients meeting these criteria were included.

### Study variables

In the analysis, changes in disease activity, HR-QoL scores and PsAID scores from baseline to Visits 3 or 4. The PsAID score ranges between 0 and 10 on the basis of responses to 12 questions, with 10 representing the worst outcome for PsA patients^23^. To evaluate the treatment effect and impact of both doses (150 mg and 300 mg), PsAID scores were divided into four categories: remission (< 2), low (≥ 2 & ≤ 3), moderate (> 3 & < 6) and high (≥ 6). Effective treatment was considered a ≥ 3-point change in the PsAID score. A t-test was performed to determine whether there were significant differences between the 150 mg and 300 mg groups. The PsAID score was selected as the primary outcome measure because of the high completeness of the data.

### Causal Machine Learning model

Causal ML encompasses a class of methods that integrate statistical techniques with ML algorithms to infer causal relationships from observational data. These models enable researchers to estimate the potential outcomes of different interventions, such as a dose adjustment, on disease activity and HR-QoL.

In the present study, the treatment effect for each patient with active PsA was estimated by comparing two potential outcomes: one under 300 mg dose treatment and another under 150 mg dose treatment. However, the individual treatment effect could not be directly observed, as each patient could be assigned to only one treatment condition.

To address these challenges, we applied the double ML framework to estimate the conditional average treatment effect (CATE), as illustrated in Figure 8.

**Figure 8:**
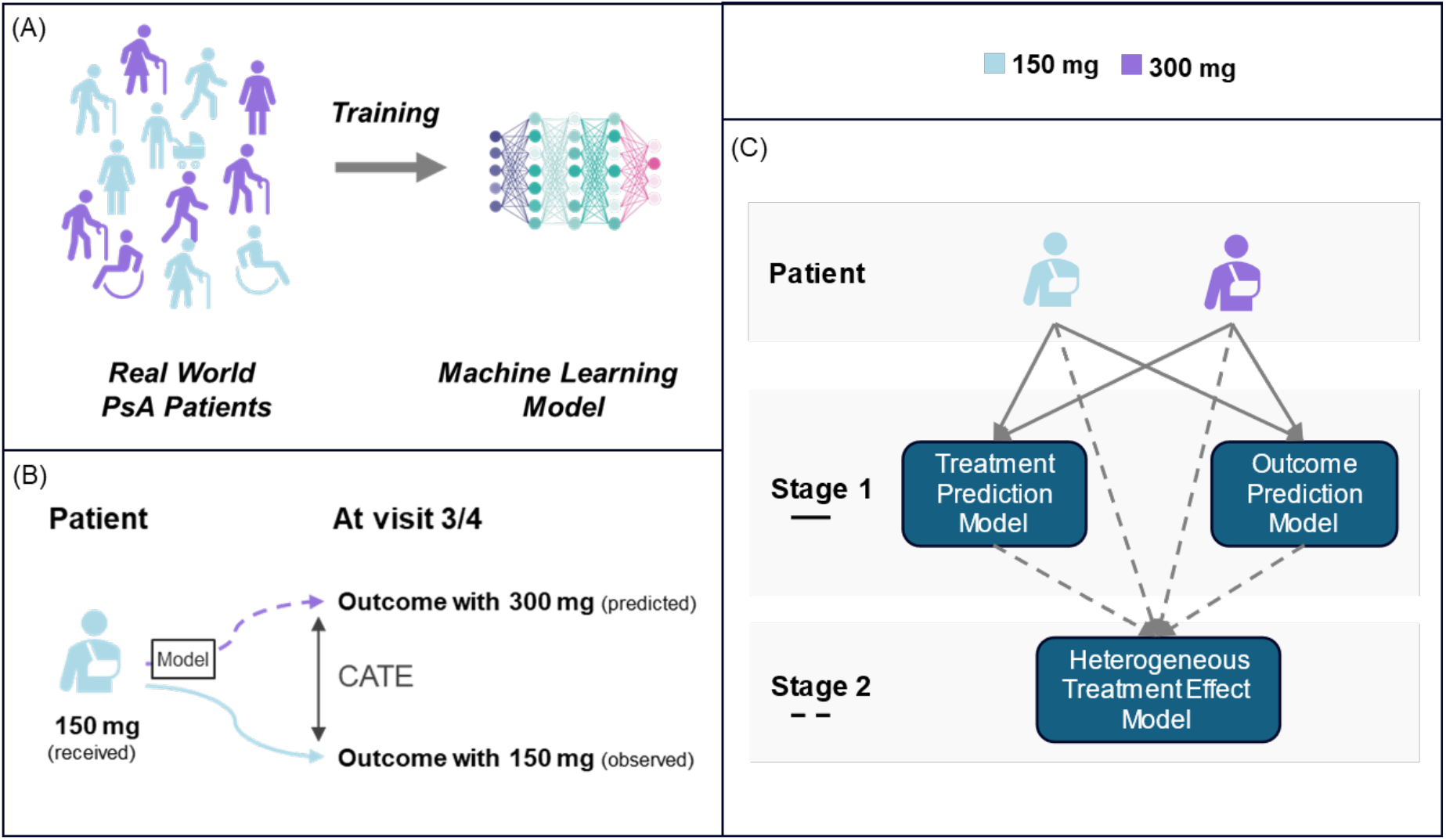
Overview of the methodology used to evaluate the effects of varying dose levels. (**a**) Development of the causal machine learning model. (**b**) Application of the model to data from patients with an initial dose of 150 mg to predict the Visit 3/4 outcome for a hypothetical initial dose of 300 mg. The same principle applied to the case of a 300 mg starting dose. (**c**) Two-stage causal inference framework (referred to as double machine learning). CATE, conditional average treatment effect

In causal inference, CATE represents the expected treatment effect for a specific subgroup or individual given their observed characteristics. In contrast, the average treatment effect (ATE) is the overall expected difference in outcomes between the two treatment conditions across the entire population. While various causal inference methods exist for different problem settings, we chose the double ML framework because of its robustness in handling high-dimensional data and its ability to provide unbiased estimates of treatment effects. The aim of our model is to quantify the effect of treatment escalation at both the individual and subgroup levels.

For this analysis, we utilized the *CausalAnalysis* class from EconML, a Python package developed by Microsoft Research that provides ML techniques for estimating individual treatment effects from observational or experimental data^24^. EconML offers a suite of causal inference tools, including double ML. These tools allow users to integrate their ML models into the framework, enabling the analysis of features responsible for causality. In addition to identifying causal factors, EconML can be leveraged to analyse and interpret appropriate interventions to address these underlying factors.

We used either the LinearDML or the CausalForestDML class for Stage 2 of the double ML framework. The “CausalAnalysis” class handles the detailed implementation of this approach, allowing us to focus on parameters such as choosing a ‘linear’ or ‘forest’ model and setting different feature subsets as confounders to untangle the selection bias. This class also includes functions to determine causal feature importance and create policy trees to identify subgroups with varied treatment responses.

Since the counterfactual value in the heterogeneity model is a predicted value (not directly observed in the data), validating these predictions is not straightforward. A counterfactual value represents what would have happened under a different treatment scenario. As a result, there is no performance metric that can be used for testing and validating counterfactuals. However, during model fitting, the CausalAnalysis class provides k-fold cross validation to evaluate causal models on the training data.

The model selection process, which encompasses performance metrics as well as the ML optimization and validation pipeline, is outlined in the supplementary material.

### Variable selection to be considered in the model

Potential inputs to ML models included 42 variables identified in our previous study^18^, including demographic characteristics, laboratory measurements, disease activity scores, HR-QoL scores, pretreatment data, and comorbidities at baseline (Visit 1). Variables with ≥ 50% missing values were removed from the test dataset.

Five types of variables were considered in the model: Y is the target variable, T is the treatment variable, W represents confounder variables affecting both the treatment and the outcome, and X includes the remaining variables that affect only the outcome:

#### Target variable (Y)

Changes in the scores of 3 disease activity measures and 6 HR-QoL measures (between baseline and Visit 3/4) were used individually as the outcome to construct causality models. The target variables for disease activity and HR-QoL included PsAID, BDI, PhGA, SPI-II, PGA, and PASI response scores. The selection of those scores was based on the content and rate of missing data.

#### Treatment variable (T)

A single variable for 150 mg (0) or 300 mg dose (1) patients starting at baseline.

#### Confounder variables (W)

Baseline value of the target variable (Y), BMI, PASI score, number of months pretreated with TNF-α inhibitors, number of pretreatments with biologics, and presence of plaque psoriasis

#### Control variables (X)

Demographic characteristics, baseline disease activity or HR-QoL scores, and comorbidities

The final feature selection for the ML model was determined through Shapley Additive exPlanations (SHAP) analyses, literature research, and model tuning to ensure optimal performance and clinical relevance; see also the supplementary section ‘Model Selection’.

## ETHICS AND CONSENT

All patients provided written informed consent for the use of their real-life data in this non-interventional study of patients with active PsA or axSpA in routine clinical practice in Germany. All methods were carried out in accordance with relevant guidelines and regulations. The study was conducted in accordance with local legislation and institutional requirements and complied with Novartis and regulatory standards, ensuring that the rights, safety, and well-being of participants were protected, consistent with the principles of the Declaration of Helsinki.

As ethics committees in Germany do not provide formal approval for non-interventional studies, a voluntary ethics consultation was conducted with the Ethics Committee of the University of Würzburg. Advice was obtained on all aspects of the study, including handling of patient data and study procedures, and all recommendations were fully implemented.

## DATA AVAILABILITY

The data that support the findings of this study are available from the corresponding author upon reasonable request, for academic purposes and in accordance with the General Data Protection Regulation (GDPR).

## CODE AVAILABILITY

The code used for causal inference modelling is available upon reasonable request for academic purposes.

## AUTHOR CONTRIBUTIONS

BG designed the study, FK prepared the data, TG performed the analysis. UK, TG, JBJ, FK, DP and BG interpreted the data. BG, TG and UK wrote the manuscript with input from JBJ, FK and DP. UK, TG, JBJ, FK, DP and BG provided critical revisions. UK, TG, JBJ, FK, DP and BG have read and approved of the final manuscript.

## ACKNOWLEDGEMENTS

We would like to thank Sean Wang for early experiments and fruitful discussions.

## FUNDING

This study was funded by Novartis Pharma GmbH.

## COMPETING INTERESTS

UK has received grant and research support and consultancy fees from AbbVie, Amgen, Biocad, Biogen, BMS, Chugai, Eli Lilly, Fresenius, Gilead, Grünenthal, GSK, Hexal, Janssen, MSD, Novartis, Onkowissen.de, Pfizer, Roche, UCB and Viatris;

TG, FK, DP and BG are full-time employees of Novartis Pharma GmbH; BG is an Strategic Advisory Board Member at Fraunhofer IZI-BB..

JBJ has no declarations in the past 36 months.

## SUPPLEMENTARY MATERIAL

### METHODS

#### Variables Construction

For this analysis MOS data was aggregated into a single variable by averaging the raw item responses of 12 items from the MOS Sleep Scale. For 10 items, higher values indicate better sleep quality. One item indicates better sleep with lower values but was not reversed for calculation. Another item, which measures average hours of sleep per night, is interpreted in a u-shaped manner, if 8 hours is considered optimal. Although this approach is not perfectly accurate, it provides a straightforward and efficient way to assess overall sleep quality.

Additionally, a variable for Patient Global Assessment (PGA) was derived by averaging the responses on the three VAS dimensions: Global Disease Activity, Global Pain and Nightly Pain.

Weighted Joint Count (WJC) was calculated as a weighted average of Tender Joint Count 68 (TJC68) and Swollen Joint Count 66 (SJC66) as follows: WJC = ⅔ * TJC68 + ⅓ * SJC66. The weights were derived from the DAS28 (CRP) formula DAS28 (CRP) = 0.56*√(TJC28) + 0.28*√(SJC28) + 0.014*GH + 0.36*ln(CRP+1) + 0.96 [Wells et al. 2009] in which the coefficient for TJC28 = 0.56 is twice the coefficient for SJC28 = 0.28.

#### Causal analysis

Using causal analysis, researchers can evaluate treatment effect heterogeneity which, in turn, enhances their comprehension of the factors influencing treatment outcomes. Additionally, it may facilitate the identification of subgroups that exhibit better response to the treatment [Kent et al 2020].

Estimating the treatment effect using observational data provides an alternative method for replicating randomized control trials (RCTs) and determining causation. However, observational studies do not have the advantage of random assignment, which makes it crucial to navigate the complexities of confounding variables and selection bias to make inferences about causation. Causal analysis can help address these challenges by providing a structured framework for untangling causal relationships from observed correlations [Ling Y et al 2023]. In most scenarios, a study population demonstrates heterogeneity, which implies that there are variations in individual characteristics that could potentially influence the effects of a treatment. In simple terms, the impact of a treatment can vary significantly within specific subgroups, deviating from the overall average treatment effect. These variations in the direction and magnitude of treatments for individuals can be explained by causal mechanisms [Yao Y, et al. 2021].

One important concept for causal analysis is unconfoundedness (that we have tracked all relevant confounders). A confounding variable (or confounder), is a factor that affects both the treatment and the outcome, leading to a misleading association. Many causal algorithms (such as the ‘Double Machine Learning’ models used in our study) are under the assumption of unconfoundedness. Although it is often impossible to measure all confounders in practice, they are not a problem if we know and track them. The selection of confounders in this study was primarily based on clinical expertise, but various combinations of covariates were also experimented with as potential confounders to observe their impact on the results.

The last important fact is that causal inference is an unsupervised learning technique that makes the task of validation challenging. The objective of causal inference is to comprehend the results of different courses of action. It is only when confirming the resilience of an estimate in consideration of unverified assumptions that the causal estimates can be deemed valid and unbiased. Typically, the robustness of the model is assessed using statistical methods, followed by validation through the insights of subject matter experts.

### Model Selection

#### Performance metrics

The first stage models were built based on the observed outcomes, allowing, in principle, for the measurement of performance. However, the CausalAnalysis class did not directly offer this possibility nor provide a means to influence the model-fitting process. We therefore applied a different strategy for model optimization, which is described below.

#### Machine learning optimization and validation pipeline

To fine-tune the model using the CausalAnalysis class, we adjusted several parameters to optimize performance. The parameters and their roles are as follows:

*heterogeneity_model*:This parameter determines the type of model used for the final heterogeneous treatment effect (stage 2). We considered two options: ‘linear’ and ‘forest’. The ‘linear’ model estimates the treatment effect as a linear function of the heterogeneity features, while the ‘forest’ model uses a forest-based approach to compute the effect from these features. Although linear models demonstrated marginally superior performance on the metric described below, we ultimately chose the ‘forest’ model, as a non-linear relationship between covariates and treatment seemed to be more plausible. *feature_inds*: The feature selection for the ML model was determined through a combination of preliminary SHAP analyses, literature research, and model tuning. Initially, SHAP analysis was used to identify the most influential features, which were then cross-referenced with findings from relevant studies to ensure their significance. Finally, model tuning was performed to refine the feature set, ensuring optimal performance and relevance to the clinical context. *heterogeneity_inds*: This parameter specifies the subset of features used as control variables (X), with the remaining features treated as confounders (W). This distinction helps in accurately estimating the treatment effect by controlling for confounding variables. *cv*: This stands for cross-validation, and we used either 5 or 10 folds (k-fold cross-validation) to ensure the robustness of our model. Cross-validation helps in assessing the model’s performance and generalizability. *mc_iters*: This parameter represents the number of times to rerun the first stage models, ranging from 3 to 20 iterations. By rerunning the first stage models with new splits each time, we aim to reduce the variance of the causal model nuisances, making the nuisance estimates from the first-stage models less noisy. To guide the tuning process, we trained 10 models using the same set of 10 random numbers and averaged the p-values of the treatment variable, with a smaller average p-value being preferable. We selected the final model based on this indicator. This approach helps in mitigating the risk of overfitting by providing a more stable estimate of the model’s performance

## RESULTS

**Supplementary Figure 1:**
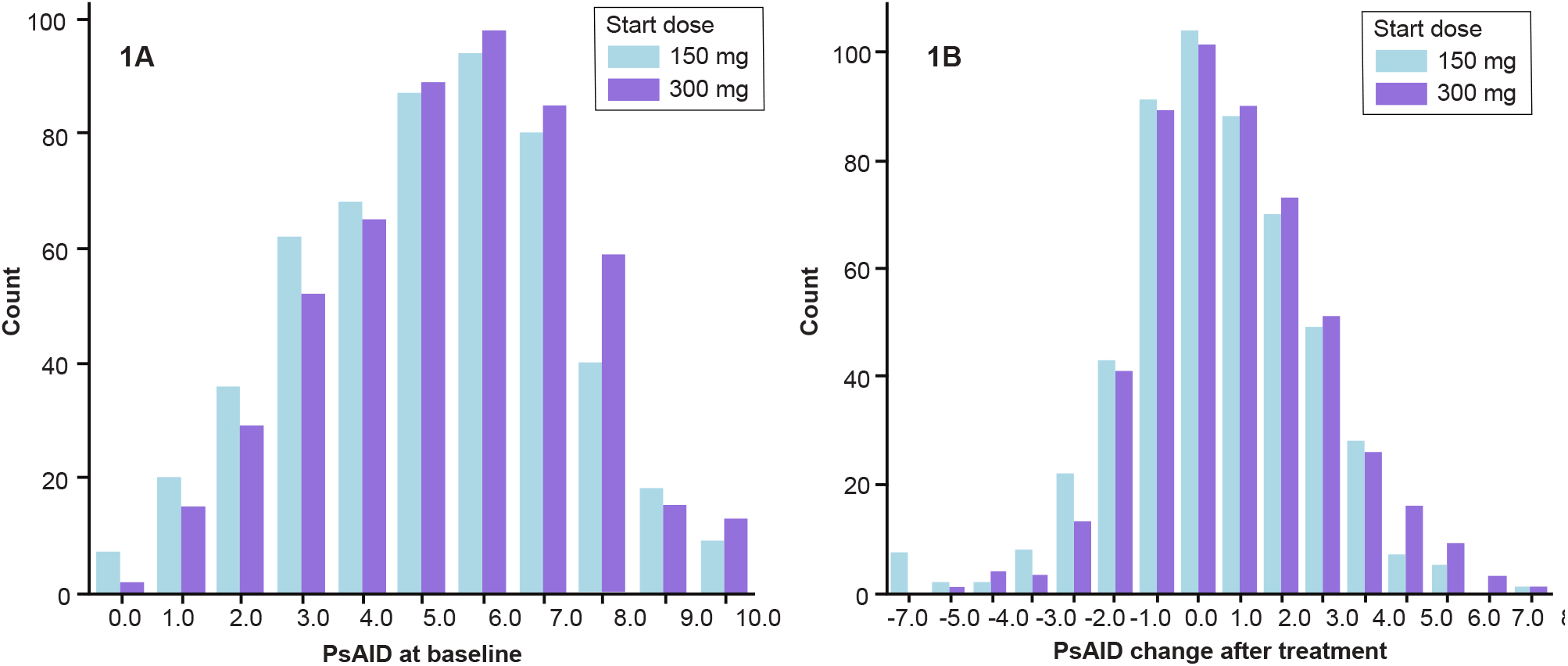
Observed PsAID distribution on baseline and PsAID change after 16-28 weeks.

**Supplementary Figure 2:**
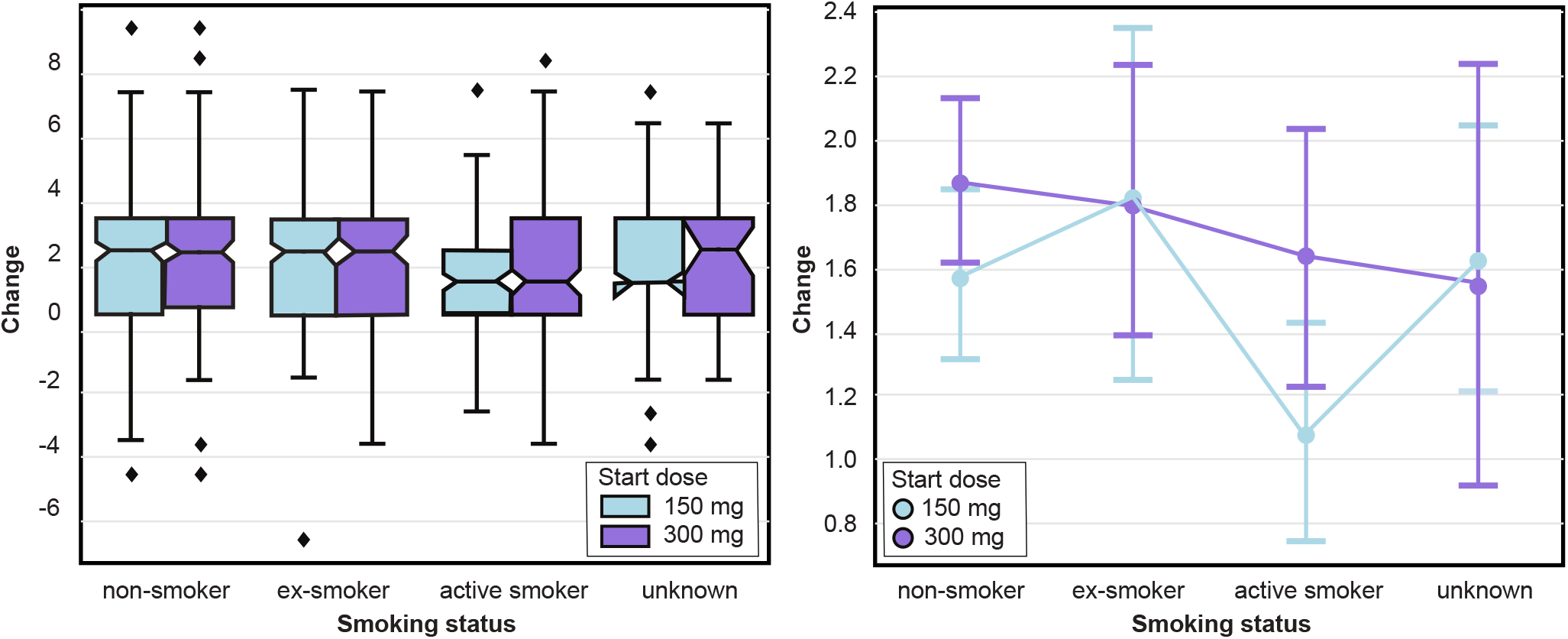
Observed PsAID treatment effect by dose as a function of status of smoking.

## Notes

### Author Declarations

The ethics committee (University of Würzburg, Germany; reference number 316/15_awb) advice was obtained and recommendations implemented.

